# Investigating shared genetic architecture between pigmentation genetics and Parkinson’s Disease

**DOI:** 10.1101/2025.04.30.25326753

**Authors:** Cristina L Abbatangelo, Brendan Newton, Frank R Wendt, Esteban J Parra

**Affiliations:** Department of Anthropology, Faculty of Arts and Science, University of Toronto, Toronto, ON, Canada; Genetics and Genome Biology Program, The Hospital for Sick Children, Toronto, ON, Canada; Institute of Medical Science, Temerty Faculty of Medicine, University of Toronto, Toronto ON, Canada; Regeneron Genetics Center, Regeneron Pharmaceuticals, Tarrytown, NY, USA; Department of Anthropology, University of Toronto Mississauga, Mississauga, ON, Canada

## Abstract

Peripheral melanin and neuromelanin share a common biosynthetic initiation. Peripheral melanin (eumelanin and pheomelanin) is cyclically produced and degraded, while neuromelanin accumulates in dopaminergic neurons over time. Neurons containing excess neuromelanin (e.g., substantia nigra) exhibit increased degeneration in Parkinson’s patients, suggesting a potential genetic interplay between pigmentation pathways and Parkinson’s Disease (PD). We used linkage disequilibrium score regression (LDSC), polygenic risk score (PRS) analysis, Mendelian Randomization (MR), and multi-trait association analysis to examine shared genetic architecture between PD and nine pigmentation-related traits (basal cell carcinoma, brown hair, melanoma, nevi, red hair, skin colour, tanning response, vitiligo, vitamin D levels). PRS analyses identified limited shared genetic variation (max 0.15% for nevi), and MR analyses did not provide evidence of a causal relationship. Together, the ten-trait and pairwise multi-trait analyses identified 48 SNPs with suggestive pleiotropy, 31 of which were protein-coding and could be mapped to 22 different genes. Overall, while some genetic overlap exists, no definitive correlative or causal relationships were established. These results contribute to the broader understanding of the differing roles of melanin and neuromelanin, as well as potential implications in neurodegenerative diseases.

## Introduction

Accumulation of neuromelanin with ageing (e.g., in the substantia nigra) is a primary risk factor for Parkinson’s Disease (PD), as neurons containing excess neuromelanin have increased vulnerability to cell death.^1,2,3^ Neuromelanin represents one of three types of the pigmented polymers known as melanin, the additional types are eumelanin and pheomelanin. For all three types, biosynthesis begins with the hydroxylation of L-tyrosine to L-dopa.^4^ Peripheral melanin (eumelanin and pheomelanin), which contributes to the colourful spectrum of phenotypes observed in hair, iris, and skin pigmentation, is cyclically produced and degraded in tissues outside of the brain. In contrast, neuromelanin in dopaminergic neurons continuously accumulates over time. The role of neuromelanin is thought to be protective, as it sequesters toxic byproducts generated by the high metabolic activity of dopaminergic neurons.^5,6^ Despite the protective qualities of neuromelanin, its accumulation over time in the neurons of the substantia nigra and locus coeruleus is a hallmark of PD.

There have been many recent attempts to bridge shared observations in pigmentation and Parkinson’s genetics using melanin.^1,2,3,4,7,8,9^ In addition to pigmented neuron loss, a second line of evidence supporting the role of melanin (and melanin-encoding loci) in PD pathogenesis is the observation that individuals with light pigmentation traits (e.g., fair skin, freckles, and blonde hair) have higher incidences of PD.^10,11^ Additionally, individuals with cutaneous malignant melanoma (CMM), exhibit higher incidences of PD and the occurrence of CMM is reciprocally higher than expected among individuals with PD.^12,13,14^ Furthermore, the largest GWAS of CMM and PD conducted to date revealed a positive and significant genetic correlation between the two phenotypes, demonstrating a shared architecture.^15^ However, the mechanisms underlying this shared genetic architecture between CMM and PD, and what role melanin- and neuromelanin-encoding loci play in this relationship, remain elusive partly because associations have been difficult to study in lab settings as many study organisms, such as mice, lack neuromelanin.^16^ Computational methods offer a unique lens as they are not limited by the physiology of laboratory organisms. This study leverages *in silico* analyses to explore potential correlative and causal relationships between PD and a variety of pigmentation traits.

## Methods

### Genome-wide datasets and global genetic correlation

The largest available set of summary statistics for PD was utilized in this study,^17^ as well as publicly available summary statistics for basal cell carcinoma, brown hair, melanoma, nevi, red hair, skin colour, tanning response, vitamin D level, and vitiligo. Sample size and additional details of the GWAS analyzed in this study are outlined in **Table 1**. Linkage disequilibrium score regression (LDSC)^18^ was used to estimate genetic correlation between PD and the nine pigmentation-related phenotypes. All P-values were adjusted using a Bonferroni multiple testing correction to determine the robustness of our findings.

**Table 1.**
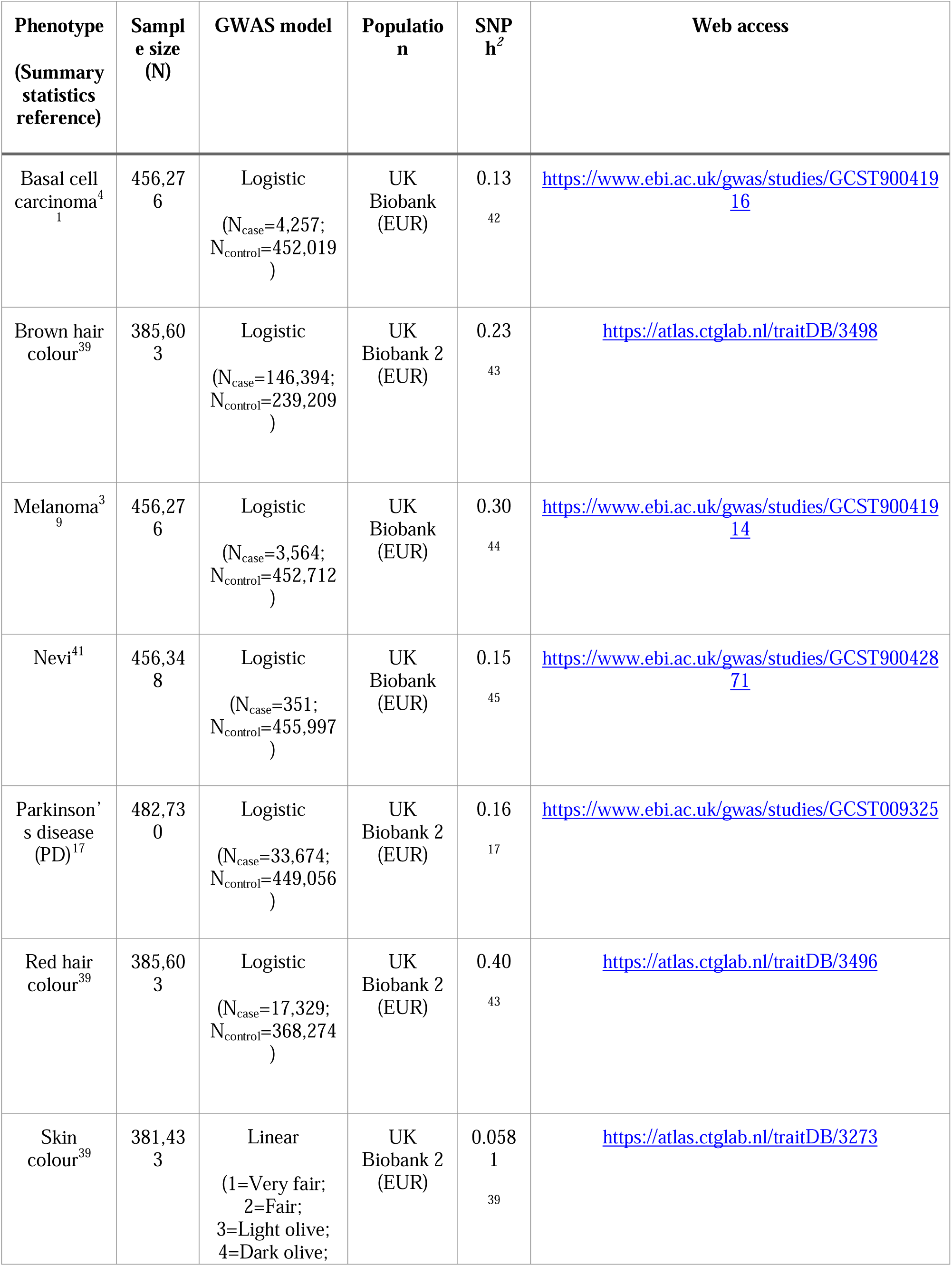
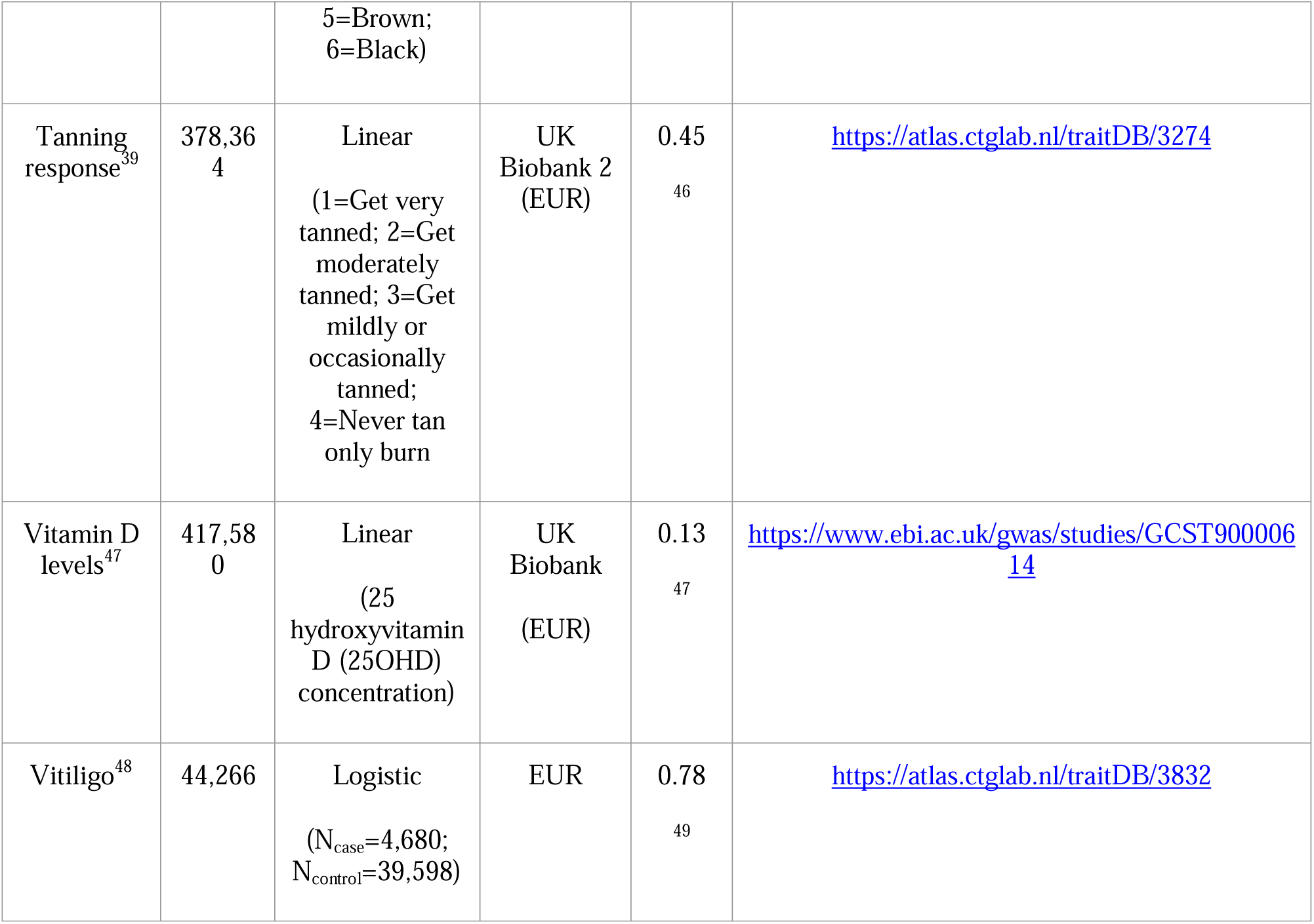
List of phenotypes used in Polygenic Risk Score (PRS) pairwise comparisons with Parkinson’s disease (PD). Sample size (N), Case and control sample size (Ncase and Ncontrol respectively), GWAS model, population ancestry and SNP heritability (SNP h^2^) for each set of summary statistics is provided.

### Determination of shared genetic architecture using PRSice

A polygenic risk score (PRS) represents the cumulative impact of genetic variants across the genome, weighted by their effect sizes on a particular phenotype.^19^ These effect sizes are typically derived from GWAS results, and only variants surpassing a specified P-value threshold are incorporated into the calculation. The calculation of PRS often involves multiple thresholds (e.g., P_T_ = 5×10^-8^, 1×10^-5^, 0.05, etc.) to ensure robust association detection. Originally applied to psychiatric conditions like schizophrenia and bipolar disorder,^20^ this methodology has been adapted for other traits to disentangle complex genetic relationships. Here we apply the first step of the program PRSice v1.25 to test for shared genetic architecture between PD and pigmentation traits.^20^ This step assesses how well a base trait can predict a target trait at different P-value thresholds. Summary statistics for PD and the 9 pigmentation traits were used as input – the analysis was conducted bidirectionally, with PD serving as the base trait for each pigmentation phenotype and then as the target trait for each pigmentation phenotype. SNPs in linkage disequilibrium were removed according to the PRSice default parameters.

### Causal inference analysis

To assess possible causal effects between PD and the pigmentation traits of interest, we conducted bidirectional two-sample MR analyses using STROBE guidelines where applicable.^21^ Two MR R packages were used: TwoSample MR v0.5.7 (https://mrcieu.github.io/TwoSampleMR/index.html)^22^ and MR-APPS (https://github.com/YangLabHKUST/MR-APSS).^23^ TwoSample MR implements MR analyses with the methods IVW, MR Egger, Weighted median, Simple mode and Weighted mode. MR-APPS is a novel MR method that accounts for pleiotropy, selection bias, population stratification and sample overlap. In a recent benchmarking analysis, MR-APPS outperformed other MR methods, producing more accurate causal effect estimates with narrower confidence intervals.^24^ With potential overlap in datasets occurring because of multiple summary statistics derived from the UK Biobank, utilization of MR-APPS in this study evaluates causal estimates independent of population structure or sample overlap between the exposure and outcome variables. PD was tested as both exposure and outcome. Note that some of the MR analyses using pigmentary traits (e.g., skin colour, tanning response, hair-colour) as outcomes can be considered negative controls as these traits are defined early in life, and we would not expect causal effects from exposures occurring later in life (e.g., PD, melanoma, basal cell carcinoma). In this case, significant results could indicate confounding due to population stratification. Exposure datasets in TwoSample MR were filtered based on the P-value threshold with the greatest variance explained (R^2^) from the corresponding PRSice analysis. Each exposure dataset was then clumped using 10,000kb windows with an r^2^=0.001 to obtain independent instruments. We used variant IDs, effect allele, other allele, effect allele frequency, effect size, standard error, and P-value for the analyses, as outlined by each program’s documentation. Sensitivity analyses detecting the presence of heterogeneity and horizontal pleiotropy were performed using various test statistics in both packages. Additionally, we cross-referenced our MR results with the iPDGC PD MR Research Portal (https://pdgenetics.shinyapps.io/MRportal/) for traits that were present in the resource (basal cell carcinoma, melanoma, tanning ability and vitiligo).^25^

### Multi-trait association analysis

We used the CPASSOC R package (version 1.01)^26^ to investigate shared genetic architecture between PD and the nine pigmentation traits used in this study. The CPASSOC package can be accessed at http://hal.case.edu/zhu-web/. To execute a CPASSOC analysis, a correlation matrix is necessary to adjust for phenotype correlations or those stemming from overlapping or related samples across different cohorts. This matrix is estimated using summary statistics derived from independent SNPs in a genome-wide association study (GWAS). The general CPASSOC approach (provided by Li and Zhu)^27^ recommends estimating these correlations using SNPs in linkage equilibrium. For datasets obtained from GWAS Catalog and GWAS Atlas, where only summary statistics are available, linkage disequilibrium (LD) patterns can be borrowed from external sources such as the 1000 Genomes Project (1KGP), which is available on the PLINK2 Resources page (https://www.cog-genomics.org/plink/2.0/resources#phase3_1kg). Continuing to follow the methods outlined in Li and Zhu’s CPASSOC general approach,^27^ the SNP selection for the correlation matrix involved LD pruning at r^2^=0.2 using PLINK2 (https://www.cog-genomics.org/plink/2.0/).^28^ SNPs with significant effects may bias correlations among summary statistics, so those with Z scores exceeding ±1.96 are excluded from the correlation matrix with the independent null set of SNPs. Then all variants are fed into S_Hom_/S_Het_ so that the calculations are correlation aware. The R code demonstrating how this is achieved can be found at https://github.com/cl-abba/Shared-Genetic-Architecture/tree/main/Pleiotropy.

CPASSOC calculates two different measures: S_Hom_ and S_Het_. The S_Hom_ method is analogous to the fixed-effect meta-analysis approach,^29^ but it incorporates adjustments for correlations in summary statistics across traits and cohorts, which may arise from related traits, overlapping datasets, or shared samples. S_Het_ is an extension of S_Hom_, permitting heterogeneity across trait effects. In this analysis, we defined pleiotropic loci as those showing a multi-trait P-value<5×10^-8^ in CPASSOC and P-values<5×10^-3^ for PD and at least one other trait. Pleiotropy was estimated between ten traits in total (basal cell carcinoma, brown hair, melanoma, nevi, PD, red hair, skin colour, tanning response, vitamin D level, and vitiligo), and also between PD and each trait separately. Setting the secondary P-value threshold (e.g., the P-value for the individual traits) too high may result in missing many true signals, and conversely, setting it too low may result in false positives, so we report the results obtained with a more stringent threshold (P<5×10^-3^, based on 0.05/number of traits assessed). The SNP2GENE function in FUMA (https://fuma.ctglab.nl/)^30^ was used to identify lead SNPs in each analysis, and SNPnexus was used to identify protein-coding variants and overlapping genes.^31,32,33,34,35^

### Gene enrichment and characterization of shared loci

Pleiotropic SNPs identified in the ten-trait CPASSOC S_Het_ runs were input as a batch query to SNPnexus.^31,32,33,34,35^ SNPs which directly overlapped protein coding genes were further explored using gene-set enrichment analysis with ShinyGO v0.741 (http://bioinformatics.sdstate.edu/go74/).^36^ Hierarchical clustering trees (P-value cutoff based on false discovery rate (FDR) was set to 0.05) generated in ShinyGo are reported. We also evaluated pairwise S_Het_ CPASSOC genome-wide significant SNPs using SNPnexus and ShinyGO for pairs of traits which illustrated evidence of sharing in the ten-trait CPASSOC analysis (PD and basal cell carcinoma, PD and melanoma. PD and brown hair colour, PD and red hair colour, PD and skin colour, PD and tanning ability, PD and vitiligo) in an effort to capture potential shared pathways between PD and individual pigmentation traits.

Overlapping genes corresponding to protein-coding lead SNPs were also further explored with STRING (https://string-db.org/)^37^ and Open Targets Genetics (https://genetics.opentargets.org/).^38^

## Data sharing

All summary statistics used are publicly available, and were accessed from GWAS Atlas (https://atlas.ctglab.nl/)^39^ and GWAS Catalog (https://www.ebi.ac.uk/gwas/).^40^ Scripts for each step of the analysis can be found on CLA’s GitHub (https://github.com/cl-abba/Shared-Genetic-Architecture).

## Results

### Genome-wide datasets and global genetic correlation

The highest genetic correlation was observed between skin colour and ease of tanning (r_g_ = 0.70) and the lowest between PD and skin colour (r_g_ = 0.016). PD showed no significant genetic correlations with any pigmentation phenotypes. Significant genetic correlations were observed between several pigmentation-related traits after multiple testing correction: skin colour and vitamin D levels (r_g_=0.21, SE=0.03, P_bonf_=2.00×10^-14^); tanning response and vitamin D levels (r_g_=0.25, SE=0.03, P_bonf_=1.80 ×10^-13^); tanning response and vitiligo (r_g_=0.28, SE=0.06, P_bonf_=1.19×10^-4^); skin colour and tanning response (r_g_=0.70, SE=0.17, P_bonf_=2.10×10^-3^); and basal cell carcinoma and vitiligo (r_g_=-0.37, SE=0.10, P_bonf_=1.29×10^-2^) (**Figure 1**).

**Figure 1.**
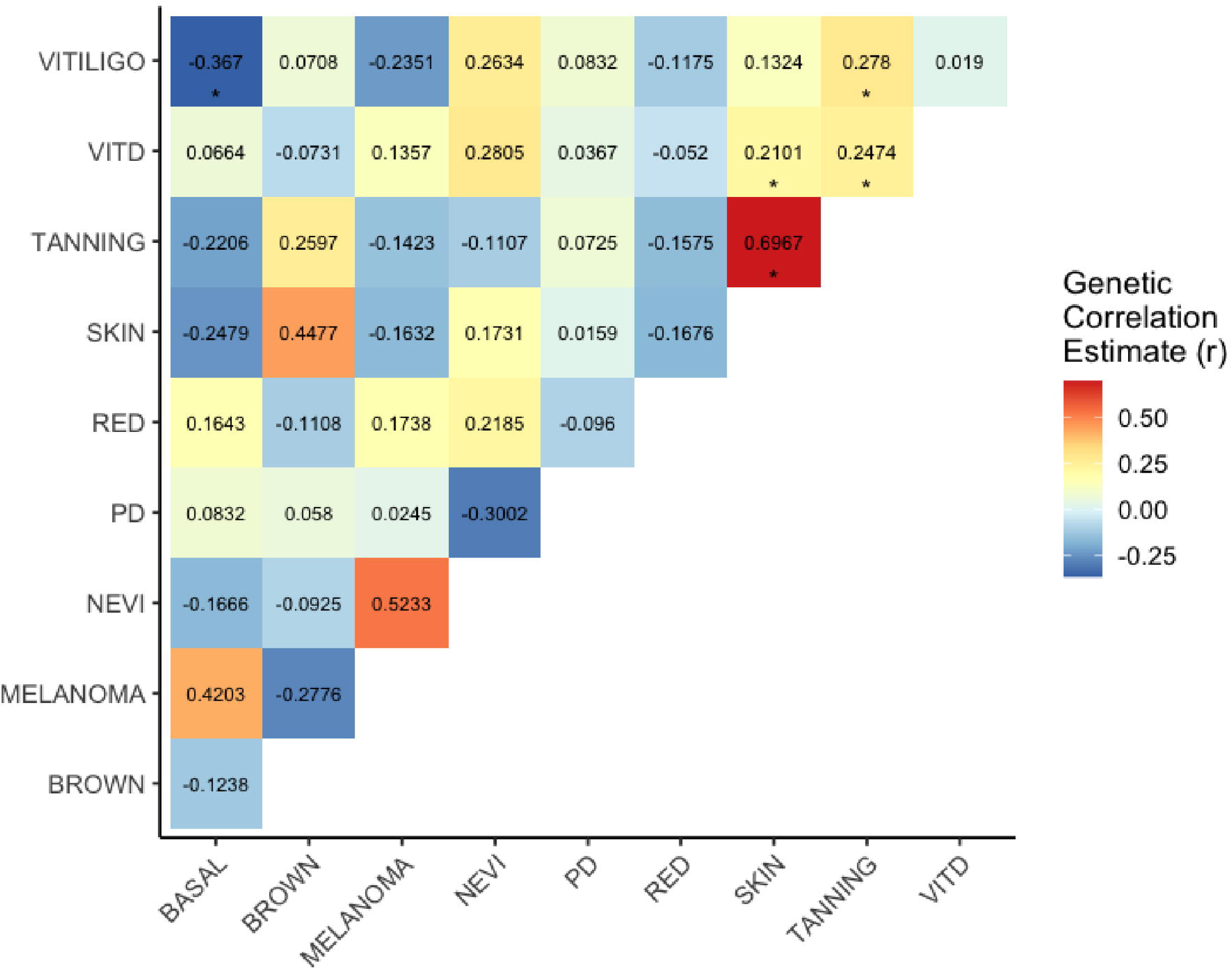
Heatmap of genetic correlations between PD and pigmentation phenotypes. P-values less than 0.05 after Bonferroni multiple testing correction are denoted by an asterisk (*).

### Determination of shared genetic architecture using PRSice

Bidirectional analyses using *PRSice* revealed some genetic overlap between trait pairs. Six phenotypes illustrated bidirectional significance with respect to PD: basal cell carcinoma, melanoma, nevus, red hair colour, tanning response, and vitamin D levels, with P-values<0.001 in both directions (significant relationship both when PD was the base phenotype and the target phenotype) although the amount of variance explained is very low (**Supplementary Table 1**). Brown hair colour showed significance only when PD was the base (P_T_=1×10^-4^).

**Supplementary** Figure 1 illustrates an example of basal cell carcinoma (base) and PD (target) to demonstrate how the P-value threshold explaining the largest percentage of variation in PD is determined. We find significant evidence that basal cell carcinoma predicts PD, with the most predictive P-value threshold of P_T_=1×10^-4^ explaining 0.55% of the variation in PD. This P-value threshold was therefore used for instrument inclusion in downstream MR steps (based on 2,634 SNPs). All P-value thresholds and the amount of variance explained are summarized in **Supplementary Table 1**. Skin colour exhibited no significant P-values and was therefore not included in the downstream MR analysis.

### Causal inference analysis

In our MR analysis, we found no clear evidence of a causal relationship of any of the pigmentation-related traits with PD. Although some of the results were nominally significant for some of the MR methods, they did not reach significance after correction for multiple testing, and the results using different approaches were inconsistent. Similar results were observed when considering PD as exposure and the pigmentation related traits as outcome. MR results are summarized in **Supplementary Table 2** and **Supplementary** Figures 2 and 3. These trends were corroborated by the iPDGC PD MR Research portal where basal cell carcinoma, melanoma, tanning ability and vitiligo also showed no significant associations.

### Multi-trait association analysis

FUMA identified 138 lead SNPs from the ten-trait multi-trait association based on the S_Hom_ statistic and 628 based on the S_Het_ statistic. S_Het_ was prioritized for future analyses as it contained all regions from S_Hom_, as well as others. The Manhattan plot for the S_Het_ CPASSOC analysis is illustrated in **Figure 2** and S_Hom_ in **Supplementary** Figure 4. None of the lead SNPs in our ten-trait analysis surpassed our thresholds for being defined as truly pleiotropic across all ten traits (e.g. CPASSOC multi-trait P-value<5×10^-8^ and P-value for each individual trait<5×10^-3^). However, we observed a few markers that satisfied the two conditions established to be considered evidence of shared pleiotropy between PD and other pigmentation-related traits: 1) The SNP demonstrated genome-wide significance in the CPASSOC S_Het_ test, and 2) the SNP demonstrated a P-value<5×10^-3^ in the individual PD GWAS and at least one other trait in the analysis (**Table 2**). Lead SNPs for pairwise tests that passed the same thresholds are summarized in **Supplementary Table 3**. The traits which illustrated sharing with PD include basal cell carcinoma, hair colour, melanoma, skin colour, tanning ability and vitiligo. Evidence of sharing was not observed between PD and nevi or vitamin D levels in the ten-trait analysis.

**Figure 2.**
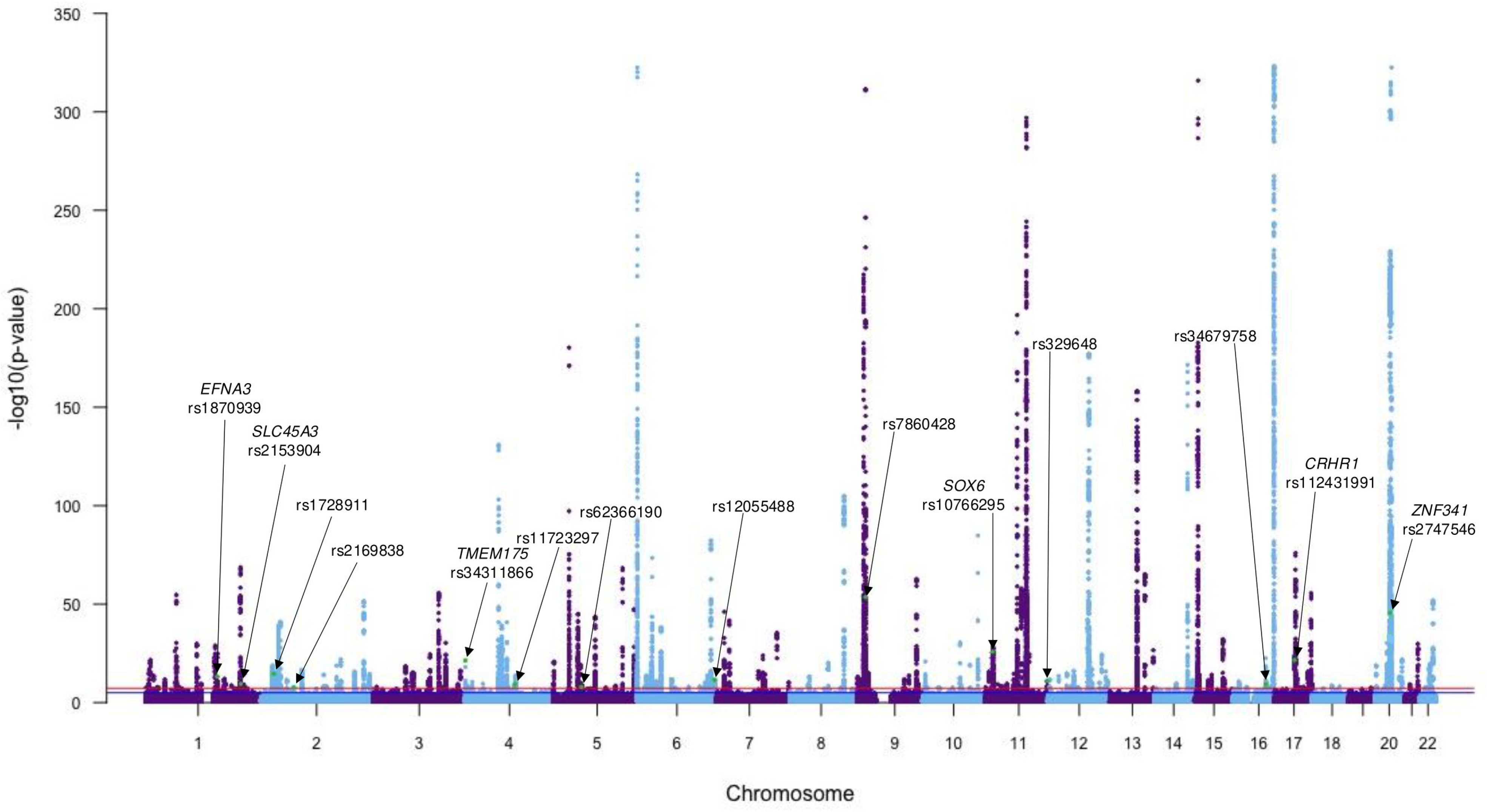
Manhattan plot depicting the results of the S_Het_ CPASSOC test (Zhu et al., 2015). Lead SNPs are highlighted in green if they satisfied the following conditions: 1) The SNP demonstrated genome-wide significance in the S_Het_ test (5×10^-8^), and 2) the SNP demonstrated a P-value<0.005 in the individual PD GWAS and at least one other trait in the analysis. A total of 14 SNPs are labelled, along with overlapped gene for protein-coding variants where applicable based on SNPnexus (Chelala et al., 2009; Dayem Ullah et al., 2012; Dayem Ullah et al., 2013; Dayem Ullah et al., 2018; Oscanoa et al., 2020).

**Table 2:**
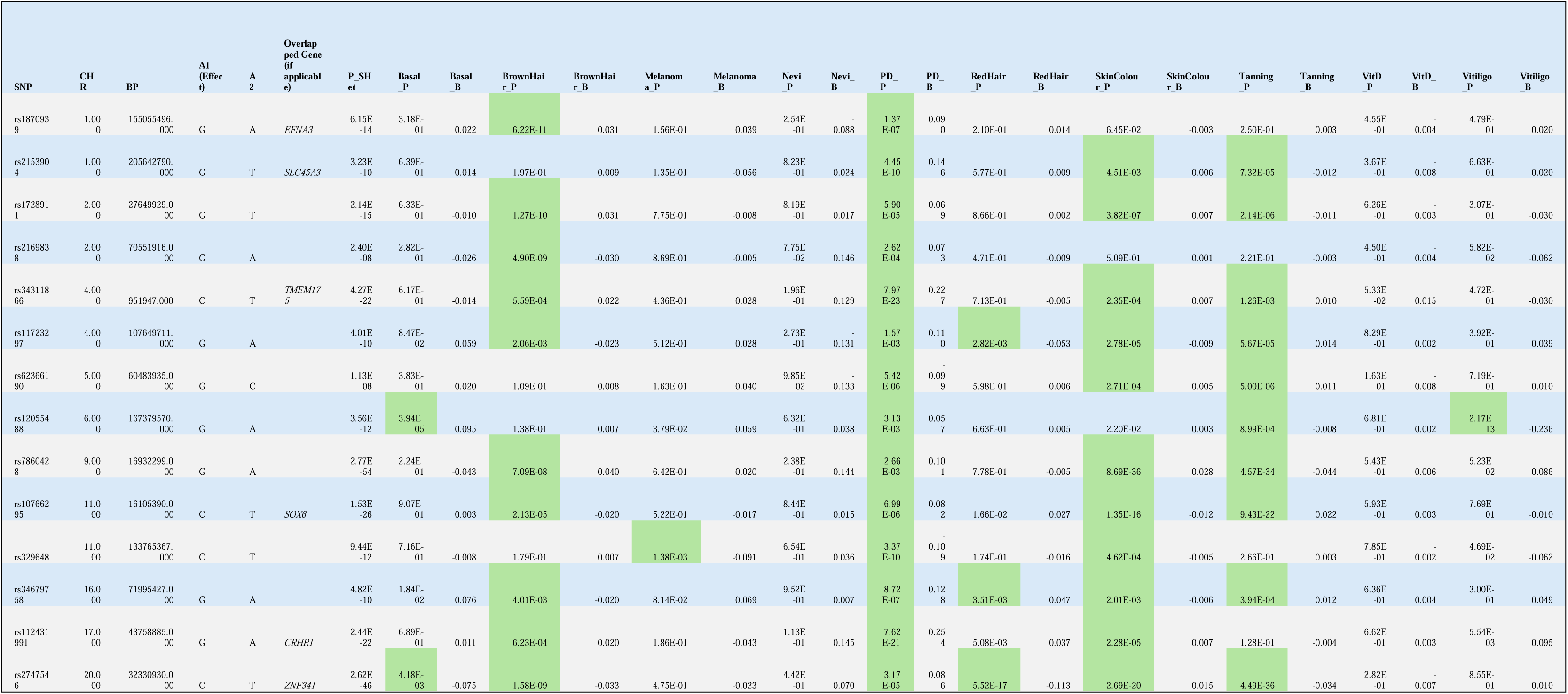
Lead SNPs from CPASSOC S_Het_ tests that demonstrated evidence of pleiotropy. SNPs are represented in the table if they satisfied the following conditions: 1) The SNP demonstrated genome-wide significance in its CPASSOC S_Het_ test (5×10^-8^), and 2) the SNP demonstrated a P-value<0.005 in the individual PD GWAS and at least one other trait in the analysis. Cells containing P-values<0.005 are highlighted in green. P-value columns formatted to Scientific notation in Excel, and Beta columns formatted to three decimal places in Excel. **SNP**: rsID identification for single nucleotide polymorphisms **CHR**: Chromosome **A1 (Effect)**: Effect allele **A2**: Non-effect allele **Overlapped Gene (if applicable)**: Variants which were identified as protein-coding have their overlapped gene listed where applicable **P_SHet**: P-value determined by S_Het_ test with CPASSOC (Zhu et al., 2015) **\*_P**: P-value associated with each individual trait GWAS **\*_B**: Beta (effect size and direction) associated with each individual GWAS **P_SHet**: Test statistic that is based on an inverse weighted meta-analysis, but maintains statistical power when heterogeneity exists

### Gene set enrichment and characterization of shared loci

**Table 3** summarizes the functional consequences of the overlapped genes corresponding to protein-coding lead SNPs from S_Het_ tests which demonstrated evidence of pleiotropy with PD. 20 of the 22 genes had evidence of expression in brain tissues according to the Genotype-Tissue Expression Portal (GTEx). **Supplementary** Figure 5 illustrates the gene network and publications enrichment for the same genes. ShinyGO results from the four pairwise CPASSOC runs SNP sets generated during the MR step are illustrated in **Supplementary** Figure 6. Across multiple analyses significant enrichment was observed in a wide variety of processes; melanin biosynthesis was commonly represented, as well as neurogenesis, metabolic processes, neuron/axon guidance, regulation of lysosomal pH and calcium mediated signaling.

**Table 3.** Protein function and expression information for overlapped genes of protein-coding lead SNPs from both the ten-trait and pairwise CPASSOC analyses. Protein function information obtained from STRING (string-db.org) and expression information obtained from GTEx Portal (https://gtexportal.org/home/) on 03/05/2025.

| **Gene Name** | **Associat**<br>**ed**<br>**Traits**<br>**with PD** | **Function** | **Expressi**<br>**on in**<br>**Brain**<br>**(Yes/No)** |
| --- | --- | --- | --- |
| *ATP8B4* | Tanning ability | Phospholipid-transporting ATPase, maintains membrane phospholipid asymmetry. | Yes (minimal) |
| *CDH13* | Brown hair colour | Calcium-dependent cell adhesion protein, may regulate neural cell growth. | Yes |
| *COL5A2* | Tanning ability | Type V collagen involved in connective tissue matrix assembly. | No |
| *CRHR1* | Brown hair colour,<br>Skin Colour,<br>Red hair colour | G-protein coupled receptor for CRH, involved in stress response and adrenal gland development. | Yes |
| *EFNA3* | Brown hair colour | Cell surface ligand for Eph receptors, involved in cell signaling. | Yes |
| *FDFT1* | Brown hair colour | Squalene synthase, involved in cholesterol biosynthesis. | Yes |
| *GCKR* | Tanning ability | Regulates glucokinase activity in response to fructose metabolites. | No |
| *GNA12* | Brown hair colour | G-protein involved in RhoA signaling, regulates transcription and phosphatase activation. | Yes |
| *GPATCH8* | Tanning ability | G-patch domain-containing protein. | Yes |
| *HCN3* | Tanning ability | Hyperpolarization-activated potassium/sodium channel. | Yes |
| *KTN1* | Tanning ability | Kinesin receptor, involved in vesicle motility. | Yes |
| *MAPT* | Red hair colour,<br>Skin colour,<br>Vitiligo | Microtubule-associated protein, involved in neuronal polarity and stability. | Yes |
| *RALY* | Basal cell carcinoma,<br>Melanoma | RNA-binding protein acting as a transcriptional cofactor for cholesterol biosynthesis. | Yes |
| *RMDN2* | Tanning ability | Regulator of microtubule dynamics. | Yes (minimal) |
| *RNASET2* | Vitiligo | Ribonuclease with activity at acidic pH, involved in RNA degradation. | Yes |
| *SETD1A* | Tanning ability | Histone-lysine N-methyltransferase, involved in transcriptional activation. | Yes |
| *SLC16A6* | Melanoma | Proton-linked monocarboxylate transporter. | Yes |
| *SLC45A3* | Skin colour,<br>Tanning ability | Glycoside-pentoside-hexuronide cation symporter transporter. | Yes (minimal) |
| *SOX6* | Brown hair colour,<br>Skin colour | Transcriptional activator involved in neurogenesis and skeleton formation. | Yes |
| *TMEM175* | Brown hair colour,<br>Skin colour,<br>Tanning ability | Organelle-specific potassium channel regulating pH and autophagy. | Yes |
| *ZC3H7B* | Brown hair colour,<br>Vitiligo | Regulator of miRNA biogenesis, binds to specific miRNA hairpins. | Yes |
| *ZNF341* | Basal cell carcinoma, Brown hair colour, Red hair colour, Skin colour, Tanning ability | Transcriptional activator of STAT3, involved in immune homeostasis. | Yes |

## Discussion

There has been increased interest in elucidating the role of melanin in diverse biological processes beyond pigmentation. Our study explored the potential genetic links between pigmentation-related traits and PD by examining their shared genetic architecture using genome correlation, PRSice, MR, and multi-trait association analyses. Despite the common biosynthetic initiation between peripheral melanin and neuromelanin, our findings do not support a correlative or causal relationship between melanin-encoding loci and PD. LDSC illustrated significant correlation among several pigmentation traits yet did not identify significant correlation between PD and any of the pigmentation-related traits. Analysis with PRSice indicated some shared genetic architecture of pigmentation-related traits with PD, however, the proportion of genetic variation in PD which could be predicted by any pigmentation trait was very small, with a maximum of 0.15% for nevi.

In a similar vein, evidence for a causal relationship based on MR analysis was weak and not replicated across different methods. An important limitation of the MR analyses in this study is the potential sample overlap between exposure and outcome datasets, particularly for traits where both sets of data are derived from the UK Biobank. Sample overlap represents a violation of the assumption of independence made in MR analyses.^50^ Methods calculated with TwoSample MR (Mr Egger, Weighted median, IVW, Simple mode, Weighted mode) are not able to correct for sample overlap, and results must therefore be considered carefully as they may be compromised by weak instrument bias or overfitting, which can cause inflation of the genetic association between the instruments (SNPs) and the outcome. For this reason, we also included analyses using MR-APPS, a recently developed approach that accounts for pleiotropy, selection bias, population stratification and sample overlap. Overall, we observed notable dispersion in beta estimates across the analyses, and none of the P-values surpassed Bonferroni-corrected thresholds for significance. Ultimately, while some initial steps are shared among melanin synthesis in the brain and peripheral epidermal tissues, MR results reveal that this shared initiation does not translate into robust causal or predictive pathways linking pigmentation genetics to PD. Future studies would benefit from the inclusion of more diverse datasets that avoid sample overlap.

Our multi-trait association analysis with CPASSOC aimed to investigate potential pleiotropic loci shared between PD and pigmentation traits. None of the markers surpassed our thresholds for being defined as truly pleiotropic across all ten traits. We identified 14 SNPs in our ten-trait analysis that demonstrated evidence of putative pleiotropy between PD and certain pigmentation-related traits, in particular hair colour, skin colour, and tanning ability. These SNPs were distributed across 10 chromosomes and included variants located near or within protein-coding genes such as *EFNA3, SLC45A3*, *TMEM175*, *SOX6*, *CRHR1* and *ZNF341*. The most significant signal was observed at rs7860428 on chromosome 9 (P_S_Het_=2.77×10^-54^), which showed associations with PD (P=2.66×10^-3^), red hair (P=8.69×10^-36^), skin colour (P=4.57×10^-34^), and brown hair (P=7.09×10^-8^). 34 additional SNPs were identified during the pairwise CPASSOC runs (**Supplementary Table 3**). While these results highlight loci with shared associations across traits, it is important to note that CPASSOC does not differentiate between true biological pleiotropy and correlated signals arising from linkage disequilibrium (LD), and some variants may tag nearby causal loci rather than exerting direct effects on multiple traits.

Many previous studies have investigated potential links between PD genetics and pigmentation traits. In a recent publication, Krainc and colleagues compiled a list of 12 genes (and associated SNPs, both rare and common) that may link skin pigmentation and PD based on functional studies and recent literature: *GCH1*, *CPNMB*, *HERC2*, *LRRK2*, *MC1R*, *OCA2*, *PRKN*, *SNCA*, *TPCN2*, *TRPM7*, *TYR*, *TYRP1*, and *VPS35*.^51^ We did not observe any evidence of significant associations between PD and any of the 12 aforementioned genes in our multi-trait analysis.

While Krainc et al.^51^ drew from functional studies and candidate gene reports, our analysis employed genome-wide approaches prioritizing statistical significance over biological hypothesis. Differences in methodology, sample composition, frequency thresholds for SNP inclusion, and the convoluted interplay of polygenicity and pleiotropy likely explain the lack of overlap. These findings highlight the complexity of translating candidate gene insights into replicable signals at the genome-wide level.

Beyond skin pigmentation, previous studies have suggested a potential link between *MC1R*, pheomelanin (the pigment responsible for red hair) and neurodegeneration, particularly in PD,^4,52^ but we did not find robust evidence of shared genetic etiology between red hair and PD. In the pairwise CPASSOC run between PD and red hair colour, only two significant loci were protein-coding (rs8080714 on chromosome 17, mapped to *SLC16A6* and rs6059655 on chromosome 20, mapped to *RALY*). This lack of association may be explained by the complex genetic underpinnings that influence both pigmentation and neurological health. While pheomelanin’s higher oxidative potential compared to eumelanin has been postulated to contribute to increased oxidative stress in dopaminergic neurons,^53^ our findings suggest that red hair colour, as a proxy for pheomelanin levels, does not significantly contribute to PD risk. Given the complicated dynamics of polygenicity and pleiotropy for both pigmentation traits and PD, it is possible that genes such as *MC1R* are involved in neurological health via pathways independent of pigmentation. Further research is needed to clarify the role of pheomelanin in PD pathogenesis, as well as to explore whether other pigmentation-related factors may influence neurodegenerative processes. Overall, our results suggest that while certain loci may contribute to both PD and pigmentation traits, the genetic overlap is quite limited.

Our gene set enrichment analysis based on the markers identified in the multi-trait association analysis with CPASSOC highlighted several biological processes of interest, though it was difficult to discern any clear pattern. Processes related to melanin biosynthesis were repeatedly represented, as well as some instances of neurogenesis, metabolic processes, neuron/axon guidance, regulation of lysosomal pH and calcium mediated signalling. Additionally, the Reference Publication (PubMed) enrichment performed with STRING identified four genes (*SOX6*, *GPATCH8*, *ZNF341* and *MAPT*) which had been previously reported in a 2020 publication, “Overlapping genetic architecture between Parkinson disease and melanoma”.^15^ Dube and colleagues identified a total of seven gene associations that passed the FDR threshold for both PD and melanoma (*GPATCH8, MYO9A*, *PIEZO1, SOX6, TRAPPC2L*, *ZNF341*, and *ZNF778*). In our study, although four genes were replicated in the multi-trait analysis, the only gene which demonstrated evidence of putative pleiotropy between PD and melanoma was *SOX6* (rs10766295). *SOX6* is a transcription factor recently implicated in the development and maintenance of substantia nigra neurons.^54^ It is primarily expressed within pigmented and tyrosine-hydroxylase positive neurons, however, in individuals with PD SOX6 expression in the substantia nigra is reduced.^54^ In mice, *SOX6* deletion leads to diminished dopamine levels and striatal innervation,^54^ consistent with PD-related pathology.^55^ A large *SOX6* deletion was also reported in a patient with developmental delay and parkinsonian features, including rest tremor.^56^ Beyond the central nervous system, *SOX6* influences gastric dopaminergic neuron development,^57^ potentially linking it to enteric nervous system dysfunction in PD. *SOX6* has also been implicated in melanoma, showing high expression in melanoma cell lines,^58^ and has been proposed as a candidate melanoma driver gene^59^ as well as a putative stem cell marker for melanoma.^60^ The observation of *SOX6* in our study reinforce the growing body of epidemiological evidence supporting a connection between CMM and PD.^12,13,14^ Although these results in the context of previous reports may suggest common regulation of gene expression both in both PD and CMM, there is still no clear indication from any of the pigmentation traits included in the analysis that melanin is the basis for this connection.

In conclusion, our findings contribute to the rising body of research aimed at elucidating the shared genetic architecture between melanin-encoding loci and PD. Although we did not identify correlative or causal links, the observed overlap in associated SNPs from multi-trait (pleiotropy) analyses provide valuable insights and suggest that shared mechanisms between PD and pigmentation may yet be at play via non-causal pathways. Advanced genomic techniques and increasing diversity in large-scale population studies offer areas of opportunity to further enhance our understanding and potentially uncover subtle, but clinically relevant, genetic links.

## Data Availability

Scripts for each step of the analysis can be found on CLA's GitHub (https://github.com/cl-abba/Shared-Genetic-Architecture). All data produced in the present study are available upon reasonable request to the authors.

## Acknowledgments

We thank Lydia M. Li (University of Toronto, Department of Applied Psychology and Human Development) for her valuable support with code troubleshooting, which contributed to the successful completion of this work.

## Authors’ Roles

CLA: Conceptualization, Formal Analysis, Writing – original draft, Writing – review and editing. BN: Formal Analysis, Writing – original draft. FW: Conceptualization, Methodology, Formal Analysis, Writing – review and editing. EJP: Methodology, Writing – review and editing, Supervision.

