## Supplementary Material for "Investigating shared genetic architecture between pigmentation genetics and Parkinson’s Disease"

Supplementary Tables

| Phenotype | PRSice P-value threshold (P_T_) (PD as Target)  (P-value (P), Variance Explained (R^2^)) | PRSice P-value threshold (P_T_) (PD as Base)  (P-value (P), Variance Explained (R^2^)) | Summary and MR directives |
| --- | --- | --- | --- |
| Basal cell carcinoma | **P_T_=1x10^-4^***  **(P=4.1x10^-60^, R^2^=5.5x10^-4^)** | **P_T_=1x10^-8^***  **(P=1.8x10^-48^, R^2^=4.5x10^-4^)** | Both basal cell carcinoma (base) → PD (target) AND PD (base) → Basal cell carcinoma (target) are significant |
| Brown hair colour | P_T_=0.0010  (P=0.071, R^2^=4.5x10^-6^) | **P_T_=1x10^-4^***  **(P=0.019, R^2^=1x10^-5^)** | Only PD (base) → Brown hair (target) is significant; run MR to see if the same pattern is observed |
| Melanoma | P_T_=0.0035  (P=0.067, R^2^=4.5 x10^-6^) | **P_T_=1x10^-4^***  **(P=0.024, R^2^=9x10^-6^)** | Only PD (base) → Melanoma (target) is significant; run MR to see if the same pattern is observed |
| Nevi | **P_T_=0.15***  **(P=7.5x10^-152^, R^2^=0.0015)** | **P_T_=0.001***  **(P=1.7x10^-227^, R^2^=0.0020)** | Both nevi (base) → PD (target) AND PD (base) → Nevi (target) are significant |
| Red hair colour | **P_T_=0.0020***  **(P=0.0060, R^2^=1.25x10^-5^)** | **P_T_=1x10^-4^***  **(P=0.020, R^2^=2.5x10^-5^)** | Both red hair (base) → PD (target) AND PD (base) → Red hair (target) are significant |
| Skin colour | P_T_=1x10^-6^  (P=0.059, R^2^=5x10^-6^) | P_T_=1x10^-4^  (R^2^=2.5x10^-5^) | No MR, PRS not significant |
| Tanning response | **P_T_=0.0040***  **(P=0.022, R^2^=8x10^-6^)** | **P_T_=0.0010***  **(P=0.002, R^2^=2.25x10^-5^)** | Both tanning response (base) → PD (target) AND PD (base) → Tanning response (target) are significant |
| Vitamin D levels | **P_T_=1x10^-6^***  **(P=1.7x10^-51^, R^2^=5x10^-4^)** | **P_T_=1x10^-6^***  **(P=7.9x10^-6^, R^2^=0.03)** | Both vitamin D levels (base) → PD (target) AND PD (base) → Vitamin D levels (target) are significant |
| Vitiligo | **P_T_=0.0050***  **(P=0.028, R^2^=7x10^-6^)** | **P_T_=1x10^-7^***  **(P=9.9x10^-271^, R^2^=4.1x10^-4^)** | Both vitiligo (base) → PD (target) AND PD (base) → Vitiligo (target) are significant |

**Supplementary Table 1**. List of phenotypes used in Polygenic Risk Score (PRS) pairwise comparisons with Parkinson’s disease (PD). The P-value threshold (PT) that captured the highest amount of shared genetic architecture (variance explained, R2) in the PRSice runs is provided for two scenarios: 1) PD as the base and 2) PD as the target. P-value thresholds that were identified as significant (P<0.05) by the PRSice output are denoted with an asterisk (*). Relationships are summarized in the final column: Green indicates a bidirectional significant relationship in PRSice, blue indicates a unidirectional significant relationship in PRSice, red indicates no significant relationships detected using PRSice. Bidirectional MR is conducted for all instances where at least one significant directional relationship was detected.

| Trait | Genetic correlation | No. of SNPs | TwoSample MR | | | | | | | MR-APPS (accounts for selection bias and population stratification/ sample overlap) |
| --- | --- | --- | --- | --- | --- | --- | --- | --- | --- | --- |
|  |  |  | *Heterogeneity* | *Pleiotropy* | *MR Egger* | *Weighted median* | *IVW* | *Simple mode* | *Weighted mode* |  |
| **PD as outcome** | | | | | | | | | | |
| Basal cell carcinoma | r_g_=0.083, P=0.27, P_bonf_=1, P_FDR_=0.39 | 68 | MR Egger Q=63, Q_DF=66, Q_P=0.58; IVW Q=64, Q_DF=67, Q_P=0.59 | Egger intercept=0.0059, SE=0.0085, P=0.49 | B=-0.088, SE=0.056, P=0.12 | **B=-0.072, SE=0.036, P=0.043*** | **B=-0.052, SE=0.023, P=0.022*** | B=-0.080, SE=0.080, P=0.33 | B=-0.069, SE=0.064, P=0.29 | B=-0.024, SE=0.048, P=0.62 |
| Brown hair colour | r_g_=0.058, P=0.24, P_bonf_=1, P_FDR_=0.39 | 415 | MR Egger Q=441, Q_DF=413, Q_P=0.16; IVW Q=445, Q_DF=414, Q_P=0.14 | Egger intercept=0.0040, SE=0.00027, P=0.068 | B=0.022, SE=0.045, P=0.64 | B=0.088, SE=0.066, P=0.18 | **B=0.079, SE=0.033, P=0.016*** | B=0.033, SE=0.16, P=0.83 | B=0.033, SE=0.049, P=0.50 | B=0.0040, SE=0.010, P=0.96 |
| Melanoma | r_g_=0.025, P=0.79, P_bonf_=1, P_FDR_=0.79 | 350 | MR Egger Q=374, Q_DF=348, Q_P=0.16; IVW Q=374, Q_DF=349, Q_P=0.17 | Egger intercept=-0.0024, SE=0.0042, P=0.58 | B=0.00020, SE=0.027, P=0.99 | **B=-0.039, SE=0.018, P=0.036*** | B=-0.013, SE=0.013, P=0.30 | B=-0.084, SE=0.060, P=0.17 | B=-0.067, SE=0.048, P=0.17 | B=-0.080, SE=0.083, P=0.33 |
| **PD as exposure** | | | | | | | | | | |
| Brown hair colour | r_g_=0.058, P=0.24, P_bonf_=1, P_FDR_=0.39 | 117 | MR Egger Q=95, DF=115, Q_P=0.91; IVW Q=96, Q_DF=116, Q_P=0.90 | Egger intercept=0.00087, SE=0.0020, P=0.68 | B=0.020, SE=0.010, P=0.050 | B=0.0060, SE=0.0068, P=0.37 | **B=0.0095, SE=0.0046, P=0.041*** | B=0.0052, SE= 0.019, P= 0.79 | B=0.0027, SE=0.014, P=0.85 | B=-0.0040, SE=0.068, P=0.95 |
| Vitiligo | r_g_=0.083, P=0.180, P_bonf_=1, P_FDR_=0.39 | 17 | MR Egger Q=26, Q_DF=16 Q_P=0.054; IVW Q=26, Q_DF=17, Q_P=0.075 | Egger intercept=0.021, SE=0.047, P=0.66 | B=-0.13, SE=0.18, P=0.48 | B=-0.049, SE=0.084, P=0.57 | B=-0.11, SE=0.063, P=0.066 | B=0.022, SE=0.15, P=0.88 | B=0.0043, SE=0.11, P=0.97 | **B=-0.39, SE=0.18, P=0.032*** |

**Supplementary Table 2**: Summary of Mendelian Randomization (MR) results for significant findings using various methods calculated with the TwoSample MR and MRAPPS R packages: Inverse-Variance Weighted (IVW), weighted median, MR-Egger, weighted mode-based estimation and corrected IVW. The first column presents the phenotypes assigned as the Exposure and Outcome. The second column provides the genetic correlation (r_g_) and associated P-values (unadjusted (P) and adjusted according to the False Discovery Rate (P_FDR_) of each pairwise relationship. The third column illustrates the number of single nucleotide polymorphisms (nSNPs) used as instrumental variables for each analysis, with outcomes related to PD and pigmentation traits. P-values less than 0.05 are denoted by an asterisk (*). Heterogeneity and pleiotropy assessments are provided alongside effect estimates for each MR method in the fourth and fifth columns. Subsequent columns provide MR results from analyses conducted using the TwoSample MR and MRAPPS packages (MRAPPS is a unified MR approach which accounts for pleiotropy and sample structure/overlap in exposure and outcome datasets).

**Notes for interpreting MR results (adapted from Davies et al. guidelines)^1^:**

*Results from MR analysis*

**nSPS**: The number of SNPs included in the analysis.

**B** (beta): The value of the causal estimate (i.e., slope coefficient, causal effect of the exposure on the outcome).

**SE**: Standard error of the causal estimate.

**P**: P-value associated with the estimate.

*Results from heterogeneity tests*

**Q**: Cochran’s Q statistic – to test the variability in the causal estimates generated for each SNP.

**Q_DF**: Cochran’s Q statistic degrees of freedom.

**Q_P**: Associated P-value with Cochran’s Q statistic.

*Results from directional horizontal pleiotropy test*

**Egger intercept**: To calculate the extent of average horizontal pleiotropy.

**SE**: Standard error associated with the intercept

**P**: P-value associated with the intercept.

| **SNP** | **CHR** | **BP** | **A1 (Effect)** | **A2** | **Overlapped Gene (if applicable)** | **P_SHet** | **Trait_P** | **Trait_B** | **PD_P** | **PD_B** |
| --- | --- | --- | --- | --- | --- | --- | --- | --- | --- | --- |
| **Basal Cell Carcinoma** | | | | | | | | | | |
| rs6059655 | 20 | 32665748 | G | A | *RALY* | 1.84E-13 | 1.45E-14 | -0.286 | 4.19E-03 | -0.104 |
| **Brown Hair Colour** | | | | | | | | | | |
| rs1870939 | 1 | 155055496 | G | A | *EFNA3* | 1.36E-16 | 6.22E-11 | 0.031 | 1.37E-07 | 0.090 |
| rs1728911 | 2 | 27649929 | G | T |  | 1.13E-12 | 1.27E-10 | 0.031 | 5.90E-05 | 0.069 |
| rs767984 | 2 | 70541938 | C | T |  | 5.98E-11 | 4.64E-09 | -0.030 | 2.25E-04 | 0.073 |
| rs78298232 | 4 | 90987657 | C | T |  | 4.48E-08 | 7.00E-05 | 0.176 | 6.07E-05 | 0.423 |
| rs34311866 | 4 | 951947 | C | T | *TMEM175* | 1.19E-23 | 0.000559 | 0.022 | 7.97E-23 | 0.227 |
| rs6897178 | 5 | 79688887 | G | A |  | 4.57E-10 | 6.50E-10 | 0.051 | 2.29E-03 | -0.089 |
| rs12213088 | 6 | 229318 | G | A |  | 1.25E-17 | 2.77E-17 | -0.080 | 4.78E-03 | -0.105 |
| rs6967664 | 7 | 2816384 | G | A | *GNA12* | 5.15E-09 | 1.05E-07 | 0.037 | 6.58E-04 | 0.113 |
| rs13276352 | 8 | 11657838 | C | T | *FDFT1* | 4.97E-08 | 0.004438 | 0.017 | 1.76E-06 | -0.112 |
| rs7848662 | 9 | 16920556 | G | T |  | 1.10E-08 | 5.10E-07 | 0.038 | 6.37E-04 | 0.116 |
| rs10766295 | 11 | 16105390 | C | T | *SOX6* | 8.65E-10 | 2.13E-05 | -0.020 | 6.99E-06 | 0.082 |
| rs2513286 | 11 | 68397316 | G | A |  | 3.47E-08 | 5.62E-07 | 0.032 | 1.41E-03 | -0.084 |
| rs2615131 | 16 | 82874016 | G | A | *CDH13* | 2.45E-08 | 0.000604 | 0.035 | 7.99E-06 | 0.201 |
| rs34679758 | 16 | 71995427 | G | A |  | 3.79E-08 | 0.004007 | -0.020 | 8.72E-07 | -0.128 |
| rs56062621 | 17 | 43798903 | G | A | *CRHR1* | 1.05E-22 | 0.000319 | 0.021 | 1.10E-20 | -0.253 |
| rs56144975 | 20 | 32356295 | G | C | *ZNF341* | 9.77E-232 | 4.05E-213 | -0.272 | 5.53E-04 | -0.131 |
| rs2281333 | 22 | 41749630 | C | T | *ZC3H7B* | 2.34E-08 | 6.03E-06 | 0.026 | 3.51E-04 | -0.072 |
| **Melanoma** | | | | | | | | | | |
| rs329648 | 11 | 133765367 | C | T |  | 2.83E-11 | 0.001383 | -0.091 | 3.37E-10 | -0.109 |
| rs8080714 | 17 | 66272177 | C | T | *SLC16A6/ARSG* | 2.20E-08 | 0.001259 | 0.091 | 1.03E-06 | -0.102 |
| rs6059655 | 20 | 32665748 | G | A | *RALY* | 1.51E-15 | 1.07E-15 | -0.370 | 4.19E-03 | -0.104 |
| **Red Hair Colour** | | | | | | | | | | |
| rs242561 | 17 | 44026548 | C | T | *MAPT* | 1.12E-19 | 0.003872 | 0.037 | 1.30E-20 | 0.251 |
| rs55938136 | 17 | 43798360 | G | A | *CRHR1* | 4.38E-08 | 0.004001 | 0.037 | 1.61E-20 | 0.250 |
| **Skin Colour** | | | | | | | | | | |
| rs34311866 | 4 | 951947 | C | T | *TMEM175* | 4.50E-24 | 0.000235 | 0.007 | 7.97E-23 | 0.227 |
| rs242561 | 17 | 44026548 | C | T | *MAPT* | 1.50E-23 | 2.85E-05 | -0.007 | 1.30E-20 | 0.251 |
| rs55938136 | 17 | 43798360 | G | A | *CRHR1* | 1.58E-23 | 3.10E-05 | 0.007 | 1.61E-20 | 0.250 |
| **Tanning Ability** | | | | | | | | | | |
| rs2153904 | 1 | 205642790 | G | T | *SLC45A3* | 1.20E-13 | 7.32E-05 | -0.012 | 4.45E-10 | 0.146 |
| rs59062514 | 1 | 155251649 | C | T | *HCN3* | 4.67E-08 | 5.72E-07 | 0.051 | 1.93E-03 | -0.241 |
| rs111271471 | 2 | 176517200 | G | A |  | 4.16E-08 | 5.39E-05 | 0.016 | 1.01E-04 | -0.147 |
| rs780094 | 2 | 27741237 | C | T | *GCKR* | 1.24E-11 | 2.02E-08 | -0.013 | 2.65E-05 | -0.073 |
| rs6716296 | 2 | 38174964 | G | A | *RMDN2* | 3.55E-09 | 4.06E-08 | -0.013 | 1.28E-03 | 0.075 |
| rs141083887 | 2 | 190031746 | C | T | *COL5A2* | 8.20E-09 | 0.000777 | 0.038 | 1.78E-06 | 0.590 |
| rs12642514 | 4 | 90629397 | C | A |  | 2.08E-11 | 0.002315 | 0.025 | 9.82E-10 | 0.320 |
| rs34311866 | 4 | 951947 | C | T | *TMEM175* | 4.65E-24 | 0.001262 | 0.010 | 7.97E-23 | 0.227 |
| rs62366190 | 5 | 60483935 | G | C |  | 1.79E-10 | 5.00E-06 | 0.011 | 5.42E-06 | -0.099 |
| rs117408811 | 11 | 97073830 | G | C |  | 4.49E-08 | 4.04E-05 | -0.034 | 1.36E-04 | 0.330 |
| rs2054095 | 11 | 16222030 | G | T | *SOX6* | 2.77E-23 | 4.00E-22 | -0.022 | 5.14E-05 | -0.079 |
| rs8007832 | 14 | 56035579 | C | T | *KTN1* | 1.82E-08 | 6.79E-07 | 0.012 | 7.87E-04 | -0.064 |
| rs7182041 | 15 | 50306583 | C | A | *ATP8B4* | 1.02E-08 | 1.00E-08 | -0.013 | 4.61E-03 | -0.056 |
| rs34679758 | 16 | 71995427 | G | A |  | 1.82E-09 | 0.000394 | 0.012 | 8.72E-07 | -0.128 |
| rs4889603 | 16 | 30982225 | G | A | *SETD1A* | 1.11E-11 | 0.000768 | 0.008 | 2.54E-09 | 0.109 |
| rs9907046 | 17 | 73025614 | C | T |  | 4.33E-08 | 1.81E-05 | -0.010 | 2.03E-04 | -0.075 |
| rs4793113 | 17 | 42580061 | C | T | *GPATCH8* | 9.58E-09 | 1.96E-07 | 0.012 | 9.71E-04 | -0.070 |
| rs12954483 | 18 | 52734527 | G | A |  | 2.74E-09 | 1.71E-07 | -0.020 | 4.19E-04 | -0.125 |
| rs2245015 | 20 | 32336577 | G | C | *ZNF341* | 2.94E-39 | 1.63E-36 | -0.034 | 2.78E-05 | 0.087 |
| **Vitiligo** | | | | | | | | | | |
| rs2757041 | 6 | 167370532 | G | C | *RNASET2* | 3.78E-15 | 1.86E-13 | -0.236 | 3.44E-03 | 0.057 |
| rs112572874 | 17 | 44072984 | G | A | *MAPT* | 1.00E-22 | 0.00338 | 0.095 | 2.93E-21 | -0.239 |
| rs5758301 | 22 | 41732389 | C | T | *ZC3H7B* | 1.14E-13 | 7.35E-11 | -0.223 | 1.10E-03 | -0.065 |

**Supplementary Table 3:** Lead SNPs from Basal cell carcinoma, Brown hair, Melanoma, Skin colour, Tanning ability and Vitiligo pairwise CPASSOC S_Het_ tests that demonstrated evidence of pleiotropy. SNPs are represented in the table if they satisfied the following conditions: 1) The SNP demonstrated genome-wide significance in its CPASSOC S_Het_ test (5x10^-8^), and 2) the SNP demonstrated a P-value<0.005 in both individual GWAS contributing to the pairwise test. P-values formatted to Scientific notation in Excel and Beta values formatted to three decimal places (Number) in Excel.

**SNP**: rsID identification for single nucleotide polymorphisms

**CHR**: Chromosome

**A1 (Effect)**: Effect allele

**A2**: Non-effect allele

**Overlapped Gene (if applicable)**: Variants which were identified as protein-coding have their overlapped gene listed where applicable

**P_SHet**: P-value determined with S_Het_ test in CPASSOC^2^

**Trait_P**: P-value associated with Brown hair, Skin colour or Tanning ability

**Trait_B**: Beta (effect size and direction) associated with Brown hair, Skin colour or Tanning ability from individual GWAS

**PD_P**: P-value associated with PD from individual GWAS

**PD_B**: Beta (effect size and direction) associated with PD from individual GWAS

**P_Shet**: Test statistic that is an extension of S_Hom_, but maintains statistical power when heterogeneity exists

Supplementary Figures

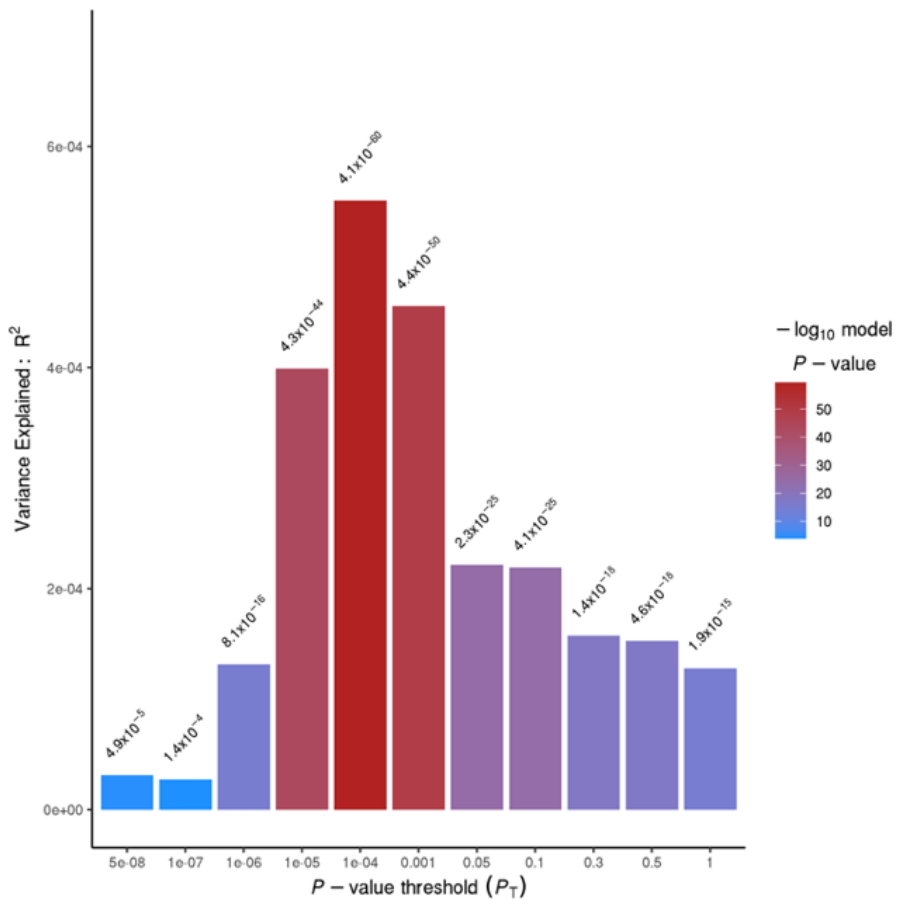
**Supplementary Figure 1.** Barplot from PRSice showing results at broad P-value thresholds for basal cell carcinoma as the base predicting PD (target). The P-value threshold (PT) accounting for the largest percentage of shared genetic architecture is P_T_=1x10^-4^.

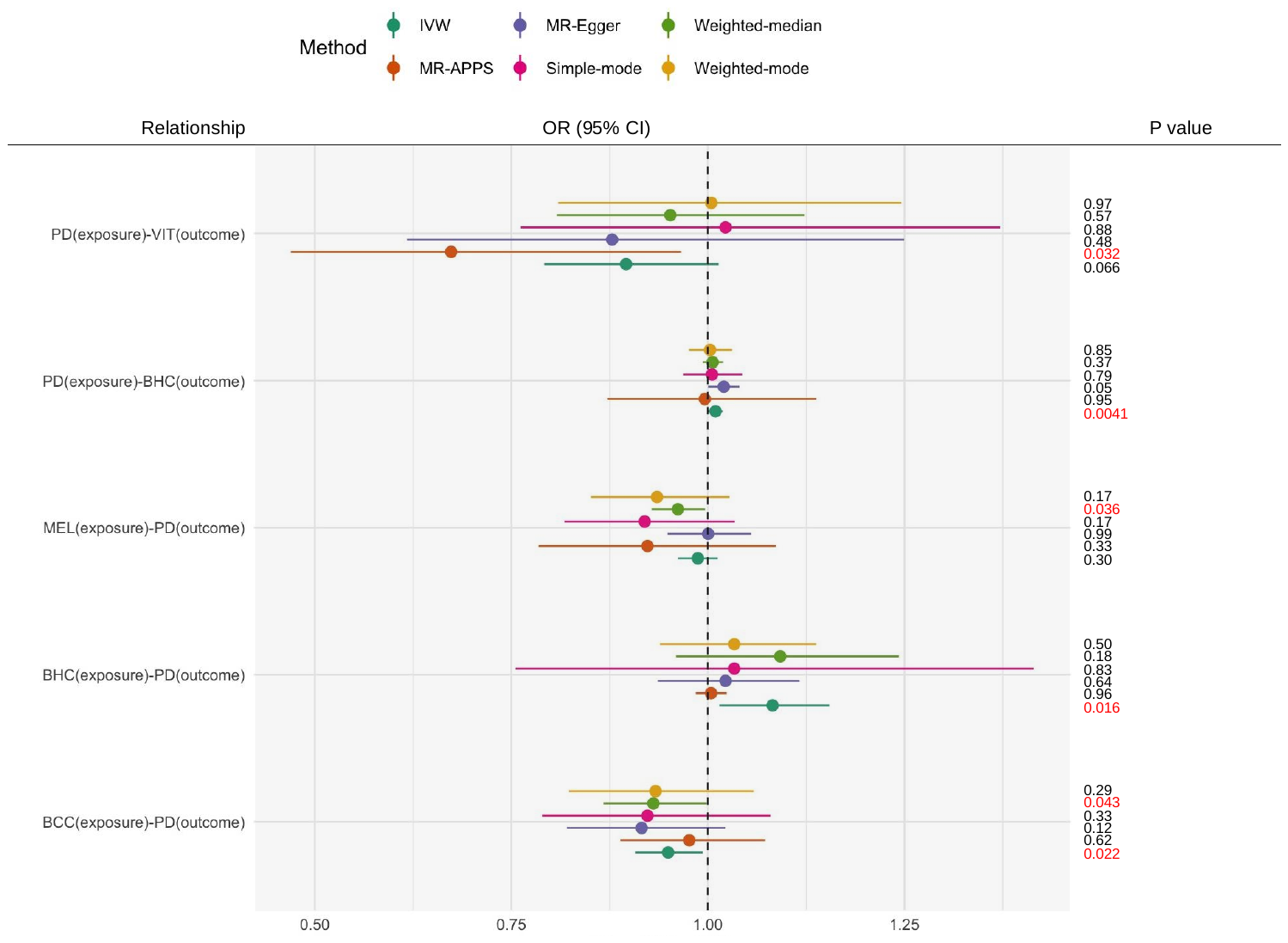

**Supplementary Figure 2.** Graphical summary of Mendelian Randomization (MR) results. None of the p-values reached significance after correction for multiple testing, and there is some evidence of heterogeneity in the results using different MR approaches. We only plot the results for pairs of traits with nominally significant findings (P < 0.05) using at least one of the methods (implemented in the TwoSample MR and MRAPPS R packages: Inverse-Variance Weighted (IVW), weighted median, MR-Egger, weighted mode-based estimation and corrected IVW. Nominally significant P-values (P<0.05) are indicated in red font.

**PD**: Parkinson’s Disease

**VIT**: Vitiligo

**BHC**: Brown hair colour

**MEL**: Melanoma

**BSC**: Basal cell carcinoma

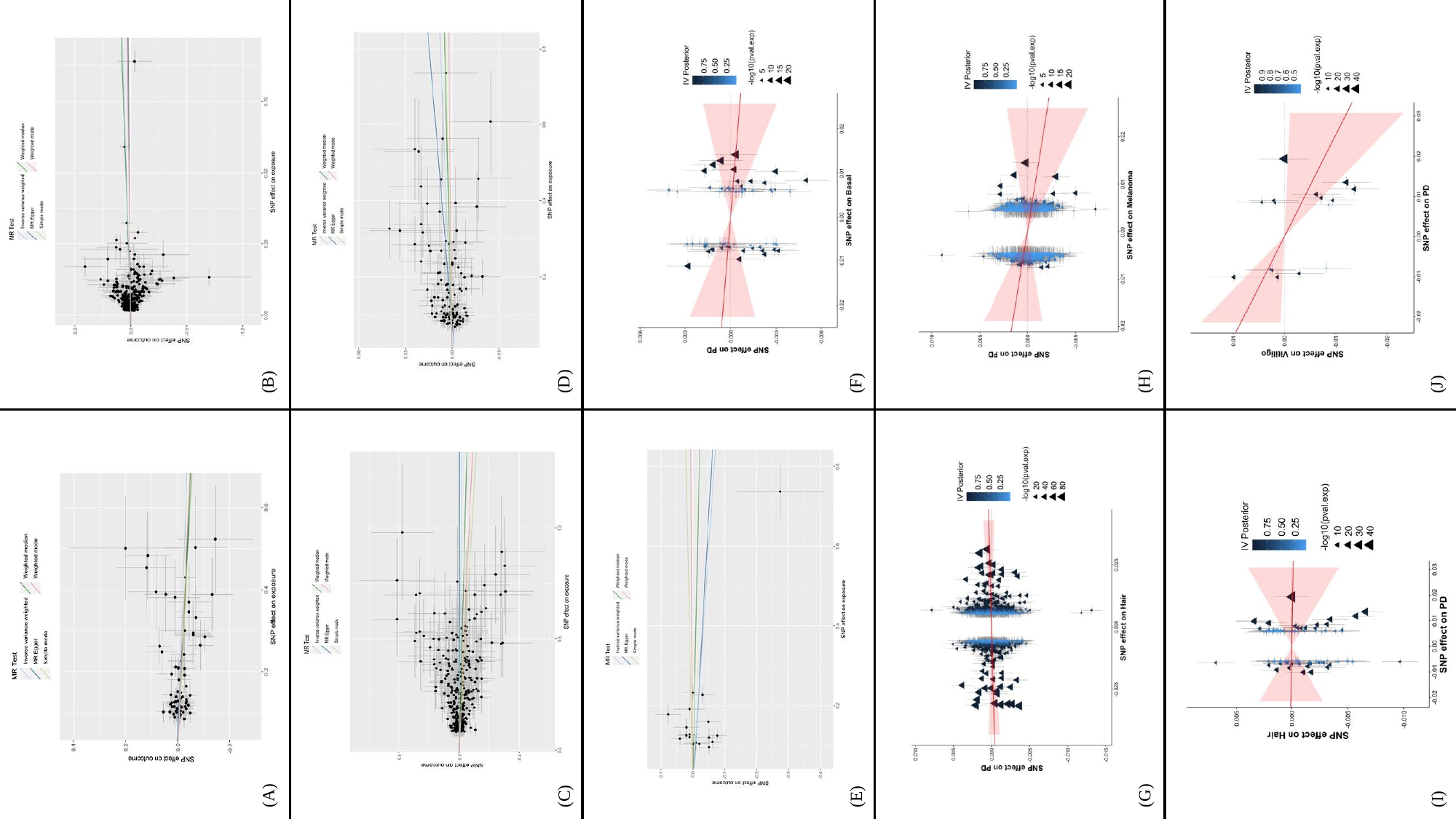

**Supplementary Figure 3.** MR plots generated using the TwoSample MR and MR-APPS R packages. Graphs **A-C** depict TwoSample MR methods where PD was set as the outcome. Graphs **D** and **E** depict Two Sample MR methods where PD was set as the exposure. Graphs **F-H** depict the MR-APPS method where PD was set as the outcome. Graphs **I** and **J** depict the MR-APPS method where PD was set as the exposure. **(A)** Basal cell carcinoma (exposure) → PD (outcome) in TwoSampleMR. **(B)** Brown hair colour (exposure) → PD (outcome) in TwoSampleMR. **(C)** Melanoma (exposure) → PD (outcome) in TwoSampleMR. **(D)** PD (exposure) → Brown hair colour (outcome) in TwoSampleMR. **(E)** PD (exposure) → Vitiligo (outcome) in TwoSampleMR. **(F)** Basal cell carcinoma (exposure) → PD (outcome) in MR-APPS. **(G)** Brown hair colour (exposure) → PD (outcome) in MR-APPS. **(H)** Melanoma (exposure) → PD (outcome) in MR-APPS. **(I)** PD (exposure) → Brown hair colour (outcome) in MR-APPS. **(J)** PD (exposure) → Vitiligo (outcome) in MR-APPS.

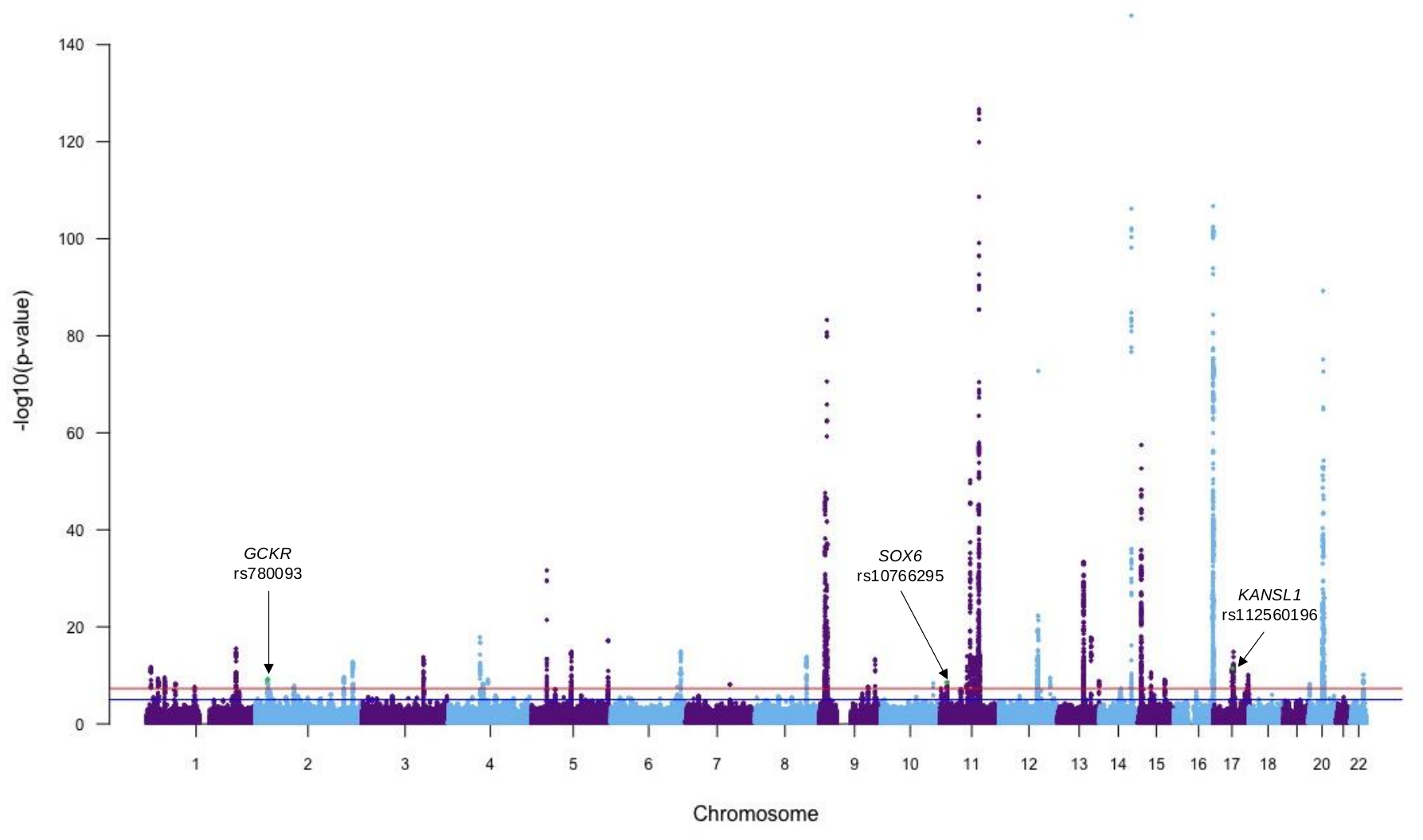

**Supplementary Figure 4**. Manhattan plot depicting the results of the S_Hom_ CPASSOC test (Zhu et al., 2015). Lead SNPs are highlighted in green if they satisfied the following conditions: 1) The SNP demonstrated genome-wide significance in the S_Hom_ (5x10^-8^), and 2) the SNP demonstrated a P-value<0.005 in the individual PD GWAS and at least one other trait in the analysis. A total of 3 SNPs are labelled, along with overlapped gene for protein-coding variants where applicable based on SNPnexus.^3,4,5,6,7^

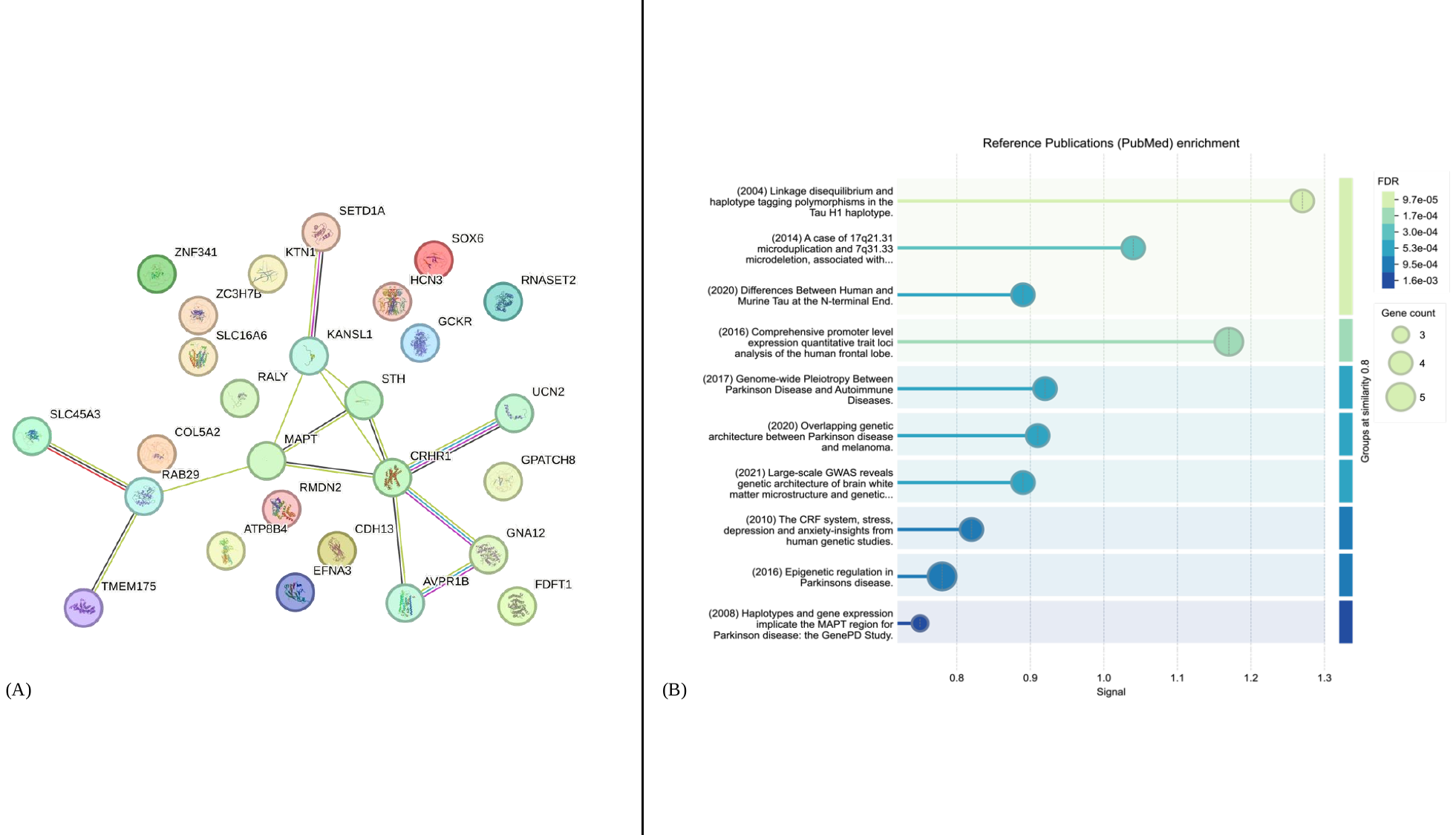

**Supplementary Figure 5**. (**A)** Network of genes directly overlapping protein-coding lead SNPs which passed both P-value CPASSOC thresholds. Each coloured circle represents a gene. Genes which demonstrate known or predicted interactions are connected with a coloured edge: Aquamarine represents known interactions from curated databases. Magenta represents known interactions which have been experimentally determined. Dark green represents predicted interactions based on gene neighbourhood. Red represents predicted interactions based on gene fusions. Dark blue represents predicted interactions based on gene co-occurrence. Light green represents connections made via textmining. Black represents connections made via co-expression. Light blue represents connections made via protein homology. (**B)** Enrichment analysis of recent publications from PubMed generated using STRING.^8^ The x-axis represents different reference publications highlighting enriched biological processes or pathways, while the y-axis indicates their significance, measured by the false discovery rate (FDR). The color gradient corresponds to FDR values, with darker shades representing more significant enrichment. The size of each bubble represents the number of genes contributing to the enrichment of a particular process. The green background highlights the most significantly enriched terms, while the blue region represents terms with relatively lower significance.

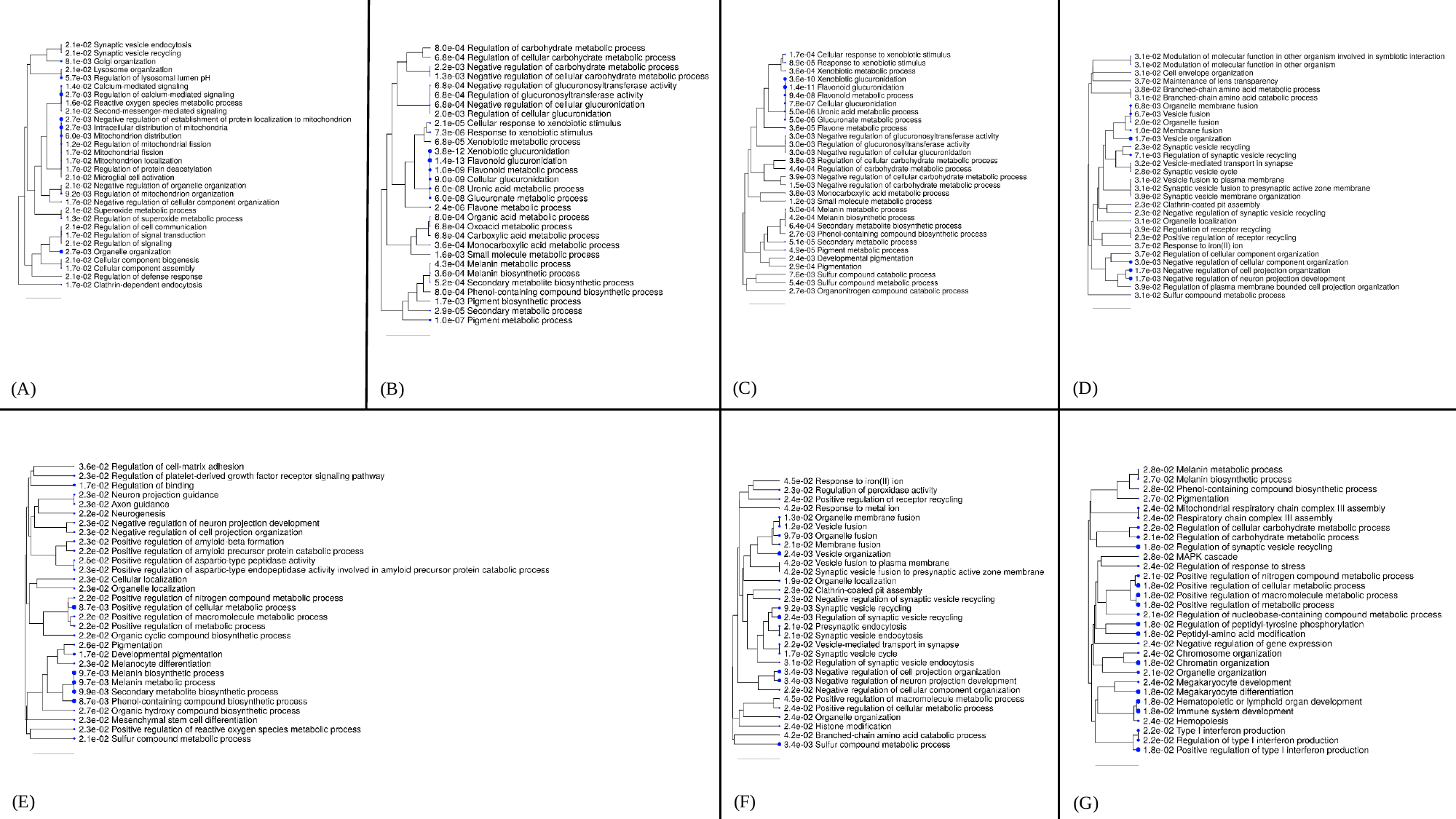

**Supplementary Figure 6.** ShinyGO results for significant pairwise CPASSOC runs. Only markers which were identified as protein-coding and had a corresponding overlapping gene were included in the ShinyGo analysis. **(A)** PD and Red hair colour. **(B)** PD and Skin colour. **(C)** PD and Tanning ability. **(D)** PD and Basal cell carcinoma. **(E)** PD and Brown hair colour. **(F)** PD and Melanoma. **(G)** PD and Vitiligo.
